# Applying Transcriptomics for an Enhanced Clinical Research Framework, Implications for an Improved Research Strategy based on an Omics Approach: A Scoping Review

**DOI:** 10.1101/2022.10.05.22280692

**Authors:** Asrar Rashid, Feras Al-Obeida, Hari Krishnan, Govind Benakatti, Wael Hafez, Joe Brierley, Benjamin Hanisch, Praveen Khilnani, Christos Koutentis, Berit S Brusletto, Mohammed Toufiq, Zain Hussain, Harish Vyas, Zainab Malik, Maike Schumacher, Rayaz Malik, Shriprasad Deshpande, Nasir Quraishi, Raziya Kadwa, Amrita Sarpal, M. Guftar Shaikh, Javed Sharief, Syed Ahmed Zaki, Rajesh Phatak, Akash Deep, Ahmed Al-Dubai, Amir Hussain

**Affiliations:** School of Computing, Edinburgh Napier University, UK; College of Technology, Zayed University, Abu Dhabi, UAE; Birmingham Children’s Hospital, Birmingham, UAE; Yas Clinic, Abu Dhabi, UAE; The National Research Centre, Egypt; Great Ormond Street Children’s Hospital, London, UK; Children’s National Hospital, Washington DC; Medanta Gururam, Delhi, India; Department of Anesthesiology, SUNY Downstate Medical Center; The Blood Cell Research Group, Department of Medical Biochemistry, Oslo University Hospital; Ullevål, Norway; The Jackson Laboratory, USA; Edinburgh Medical School, University go Edinburgh, Edinburgh, UK; Nottingham University, Nottingham, UK; College of Medicine, Mohammed Bin Rashid University of Medicine and Health Sciences, Dubai, UAE; Sheikh Khalifa Medical City, UAE; Institute of Cardiovascular Science, University of Manchester. Manchester, UK; Weill Cornell Medicine-Qatar, Doha, Qatar; Centre for Spinal Studies & Surgery, Queen’s Medical Centre. The University of Nottingham. Nottingham, UK; NMC Royal Hospital, Abu Dhabi, UAE; Sidra Medicine, Doha, Qatar; Department of Endocrinology, Royal Hospital for Children, Glasgow, UK; All India Institute of Medical Sciences Hyderabad; Pediatric Intensive Care, Burjeel Hospital, Najda, Abu Dhabi; Pediatric Intensive Care Unit, King’s College Hospital, London, United Kingdom

## Abstract

Sepsis remains a major global health issue in pediatric and adult populations, largely due to a lack of understanding of its complex pathophysiology. Despite its high mortality rate, there have been few advancements in sepsis-specific therapies over recent decades. The study aimed to investigate the potential benefits of a genome-wide transcriptomic approach to sepsis in pediatric and adult populations in reducing sepsis-related mortality and enhancing sepsis guidelines. The scoping review explored gene expression data pertinent to developing sepsis guidelines related to its definition, classification, disease severity, molecular biomarking, and benchmarking. A system-biology approach using transcriptomics was adopted to enhance the understanding of sepsis at the mRNA gene expression level. The study involved a search of the PubMed database for original research or systematic reviews that involved transcriptomic application in the context of clinical sepsis published over a ten-year period, from 2012-2022. Of the 14,048 studies retrieved, a full-text analysis was performed. Five main concepts emerged: case definition, classification, quantifying severity of sepsis, transcriptomic biomarkers, and benchmarking. Studies were categorized according to these five categories. The results showed evidence of a connection between the transcript and clinical sepsis, demonstrating that transcript-driven sepsis categorization is possible. Integrating transcriptomic data with clinical endpoints holds promise for more precise sepsis treatment. Although further exploration is needed, the methodology shows potential for disease modification.

## Introduction

Sepsis continues to be a major contributor to in-hospital mortality globally and poses a significant public health burden, particularly in pediatric populations ^1^. Consensus guidelines have been developed for managing sepsis in both adult and pediatric patients, emphasizing early recognition and intervention, aiming to standardize sepsis management practices ^2, 3^. However, challenges persist in creating protocols due to gaps in clinical evidence, as a relevant framework to understand sepsis has not been finalised. A reason why adult sepsis definitions have undergone many iterations in the past decades ^4, 5^. The pediatric surviving sepsis guidelines exemplify the impact of an unclear definition for developing guidelines feature mostly weak recommendations due to low-quality evidence ^6^. These knowledge gaps have helped identify numerous research priorities and pathophysiological questions. The prevailing view of sepsis as a clinical phenomenon has potentially limited the comprehension of its molecular underpinnings, which may hinder the identification of diagnostic and therapeutic targets, as well as the establishment of a definitive sepsis definition applicable to all age groups^7–9^.

Despite potential therapeutic breakthroughs for sepsis, such as vitamin C^10^ and activated protein-C (APC), these have shown limited success in clinical trials or meta-analyses ^11^. While successful in cancer and viral infections, immunotherapy has not gained traction in bacterial sepsis due to its affect on a multitude of molecular pathways, resulting in immune dysfunction. The slow progress in sepsis treatment emphasizes gaps in scientific knowledge ^12, 13^ and the need for early recognition, diagnosis, and resuscitation. Current laboratory protein biomarkers like CRP and Pro-calcitonin have limited predictive capabilities ^14^, and the inability to stratify sepsis patients based on biochemical and immunological profiling further complicates matters ^15 16^.

Sepsis represents a complicated challenge due to its multifactorial heterogeneity resulting in a variable disease process ^17^. Further, the transition from infection to septic shock remains poorly understood due to factors such as innate and adaptive immune mechanisms, the severity of infection, host age, adequacy of treatment, and genetic variability/susceptibility ^18^. Moreover, Genetic determinants for sepsis are only partially understood, adding to the complexity. Additionally, the quality of clinical care provided to patients with severe infections influences the disease’s natural history, with serious consequences in children ^19^.

Omic approaches, including lipidomics, metabolomics, proteomics, and transcriptomics, have been employed to understand complex disease paradigms like sepsis. Transcriptomics focuses on analyzing all RNA transcripts in a biological system, allowing a study of gene expression patterns, detection of differentially expressed genes, and description of alternative splicing events. Hasson et al. (2022) highlighted the potential of transcriptomic analysis in understanding sepsis-associated acute kidney injury and uncovering underlying pathophysiological mechanisms ^20^. Transcriptomics offers a systems-based approach to understanding biological processes ^21^ and facilitates precision medicine strategies ^22, 23^. The enhancement of in silico techniques has facilitated system-wide gene expression analyses, contributing to a burgeoning body of knowledge. Consequently, the current study seeks to explore the application of transcriptomic analysis to deepen our understanding of sepsis. This progression is the next sequence in the genetic dogma, signifying a transition from DNA to mRNA, including not only the study of protein-encoding genes (from coding regions of the DNA) but also non-coding regions that generate microRNA, circular RNA, and long noncoding RNAs. One of the key advantages of RNA-based technologies over DNA studies is the ability to reveal real-time dynamic changes to develop a temporal understanding of sepsis.

Hence a scoping review was undertaken to understand whether findings from transcriptomic studies can be implemented into clinical practice, particularly into international sepsis guidelines. As a part of this gaps in the literature were also to be identified.

Therefore a principal aim of this scoping review was to scrutinize the peer-reviewed literature spanning a decade, from 2014 to 2023, with a primary focus on applying transcriptomic findings to clinical sepsis. In order to translate facets from the research literature and interdigitate these with gene expression studies, a framework is presented (Figure 1). An enhanced framework holds potential clinical advantage, such as paving the way for guideline development, standardizing care pathways and advancing the principles of precision medicine. The framework as illustrated uses key aspects crucial in establishing evidence-based sepsis guidelines. An important objective was to discern whether transcriptomic studies can lend support for an enhanced understanding sought by peer-lead sepsis committees. This work will have important implications for research as gaps shall be highlighted with implications for guidelines and clinical practice depending on the presence of relevant research literature.

**Figure 1.**
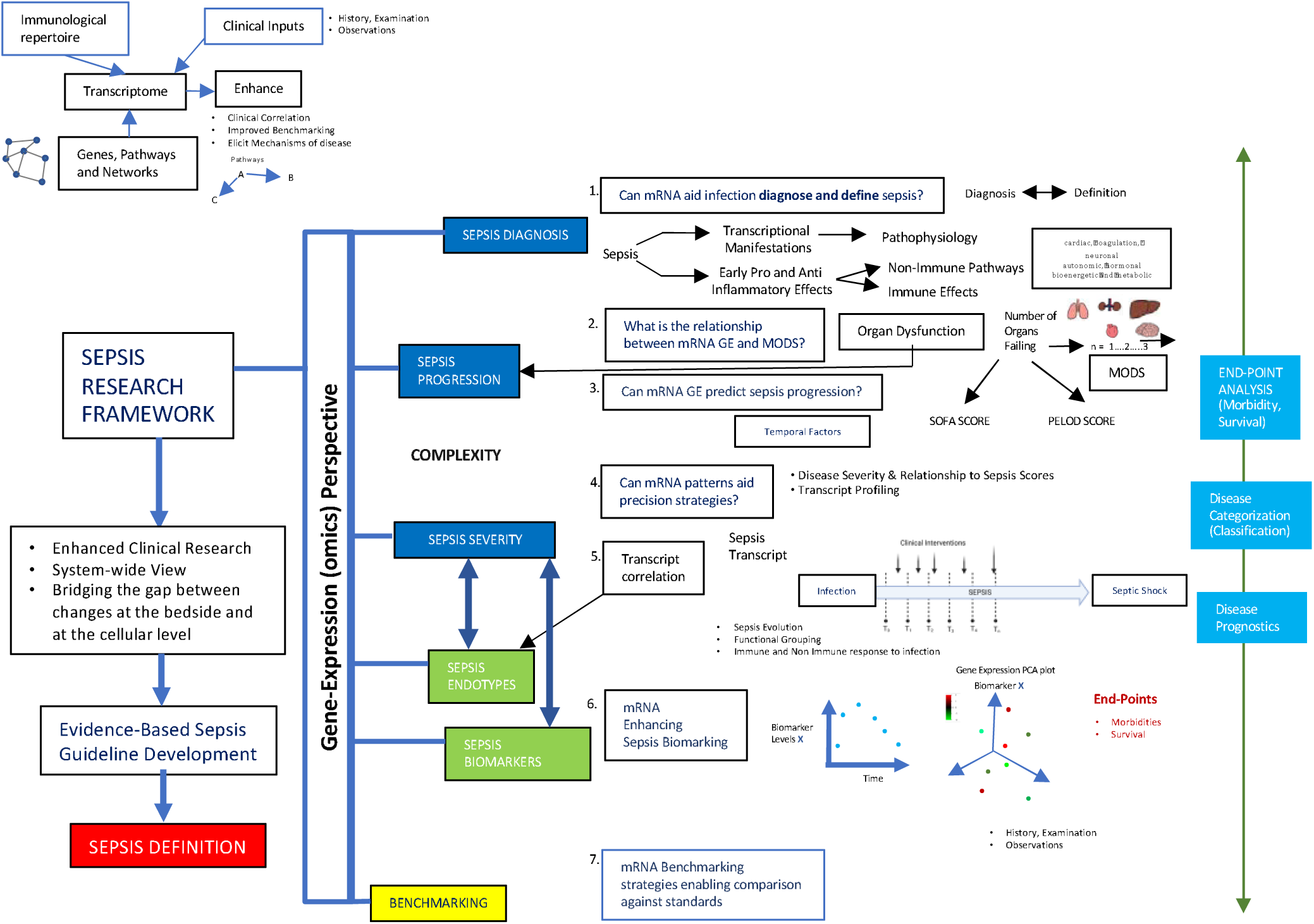
Sepsis Framework. Using transcriptomic information to support translating clinical sepsis research to the bedside. Key questions linking gene expression (GE) to the framework are shown (central boxes). These then support components of the research framework (Diagnosis/Definition, Disease Progression, Disease Severity, Biomarking, and Benchmarking). mRNA is thus vital for cellular function and consists of mRNA protein-coding and non-protein-coding RNA functions. The two facets allow mRNA to play a role in gene code translation for protein synthesis and a gene regulatory role. Essentially, mRNA is the genetic mediator guiding ribosomal protein synthesis based on information provided in the DNA. At this moment, transcriptomics aims to document gene activity by quantifying mRNA, analyzing gene expression patterns, and measuring gene levels in sepsis. Gene-to-gene connections are shown, with genes illustrated as nodes. The interconnections between the genes, then, represent the regulatory relationship. Therefore the network interactions amongst the genes form a Gene Regulatory Network (GRN). Sepsis is a heterogeneous process impacted by host factors such as age, infection timing, and pathogen-associated factors. Red and dark blue are clinical attributes; green pertains to a biological construct related to a cellular function or clinical end-point; yellow depicts a standard against which other parameters are compared. In light blue are target end-points which can be deduced from the data features, according to the generated (vector) of data-points for each patient. **1.** The diagnosis and definition of sepsis are interrelated. Changes in cellular activity can be detected with respect to mRNA GE, providing patterns indicative of a pathophysiological response indicative of sepsis. Thereby resulting in GE patterns consistent with the diagnosis and definition of sepsis. **2.** Sepsis progression is important because, if unabated, this can then progress to multi-organ dysfunction syndrome (MODS). Single-organ dysfunction in sepsis is rare, with the subsequent failure of each organ being associated with an increased risk of a poor outcome^68^. The Sepsis 3 task force concluded that the misleading model that sepsis follows a continuum from severe sepsis to shock was misleading^17^. Further, the task force concluded that the term severe sepsis was redundant. **3.** Applying transcriptomic methods to sepsis is in predicting organ dysfunction. Scoring systems exist to help quantify the degree of organ dysfunction. The sequential Organ Failure Assessment (SOFA) score is used in adult sepsis and pediatric logistic organ dysfunction (PELOD) in children. **4.** An essential aim of the transcriptomic analysis is to improve the application of clinical therapies in a more precise approach, mindful of host-pathogen complexity. The aim is for therapies to be tailored according to a specific profile or sepsis sub-type. Sepsis Subtyping may be undertaken from a gene function perspective, such as according to a distinct pathophysiological mechanism known as Endotypes. **5.** Understanding how the transcript correlated to disease severity allows the linkage of mRNA GE, a proxy of cellular function, to clinical categorization. Clinical categories of different severity levels include sepsis, severe sepsis, and septic shock. Relating the gene transcript to different levels of disease severity could allow insights into sepsis pathogenesis and provide an interpretation of the host-infection relationship. **6.** The relationship of biomarkers to gene expression is of particular interest, especially from a temporal perspective allowing the tracking of sepsis, therefore, understanding disease trends when managing patients. This can be used in a predictive capacity and to pre-empt disease progression, thereby providing information to the clinician to make management choices. **7.** The transcript may also have value in benchmarking sepsis, such as correlating to clinical variables, standards, and endpoints. Transcriptomics provides the ability to enhance endpoint analysis, aiding in disease categorization/classification and with respect to prognostication. One of the challenges in developing a clinical research framework for sepsis is that the components defining the framework may not be clear due to the lack of clarity in the original definition of sepsis. Therefore the likely overlap between components, for example, thought endotype, could allow the clustering of groups of patients; this approach may also have value as part of a biomarking strategy.

## Materials and Methods

In accordance with the PRISMA-ScR guidelines ^24, 25^, a scoping review was conducted to investigate the use of systems-biology approaches in examining gene expression and its relation to clinical sepsis. The protocol was registered on the Open Science Framework (https://osf.io/3jbv2), with the associated project osf.io/5c2wr.

### Identifying the research question

This study hypothesized that transcriptional research could support a clinical research framework based on components important for sepsis guideline design. The research question posed was: What gene expression studies can be utilized to capture relevant information for the development of sepsis guidelines concerning sepsis definition, classification, disease severity, molecular biomarkers, and benchmarking? This study incorporates gene expression investigations that span both coding and non-coding domains. The research encapsulates micro RNA studies (<200 NBP), along with the exploration of circular (Circ) RNAs and lncRNAs.

### Study Selection

A systematic review of peer-reviewed literature was undertaken from PubMed-indexed journals utilizing PubMed’s online search tool, which incorporates MEDLINE, PMC, and BookShelf databases. Pertinent literature from a 10-year period leading up to the search date, 29th June 2023 was undertaken. The primary search title terms included ’gene expression’ and ‘Sepsis’ in conjunction with one of the thematic research terms (Endotype, Biomarker, Definitions, Diagnosis, Progression, Severity, and Benchmark) (Figures 2A-G). The derived search string is shown (Supplement Table 7). At the subsequent stage, articles were filtered to include human studies in English, whilst excluding review articles, drug and vaccine studies. As the key theme ’Benchmark’ yielded zero selections, the filtration step was expanded beyond the title for a full article search. This modification was also applied to the ’Endotype’ theme to secure a more comprehensive collection of studies.

**Figure 2.**
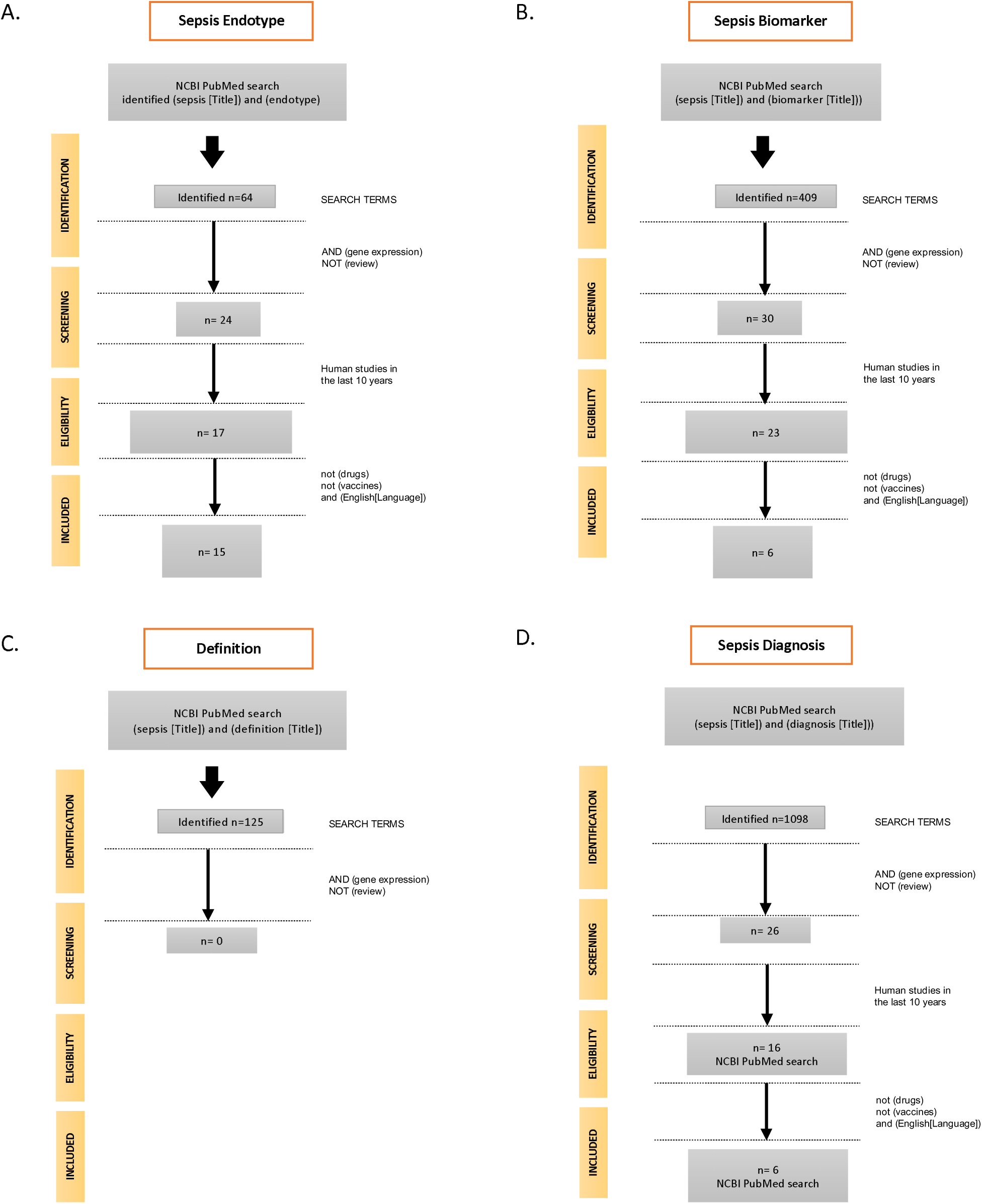
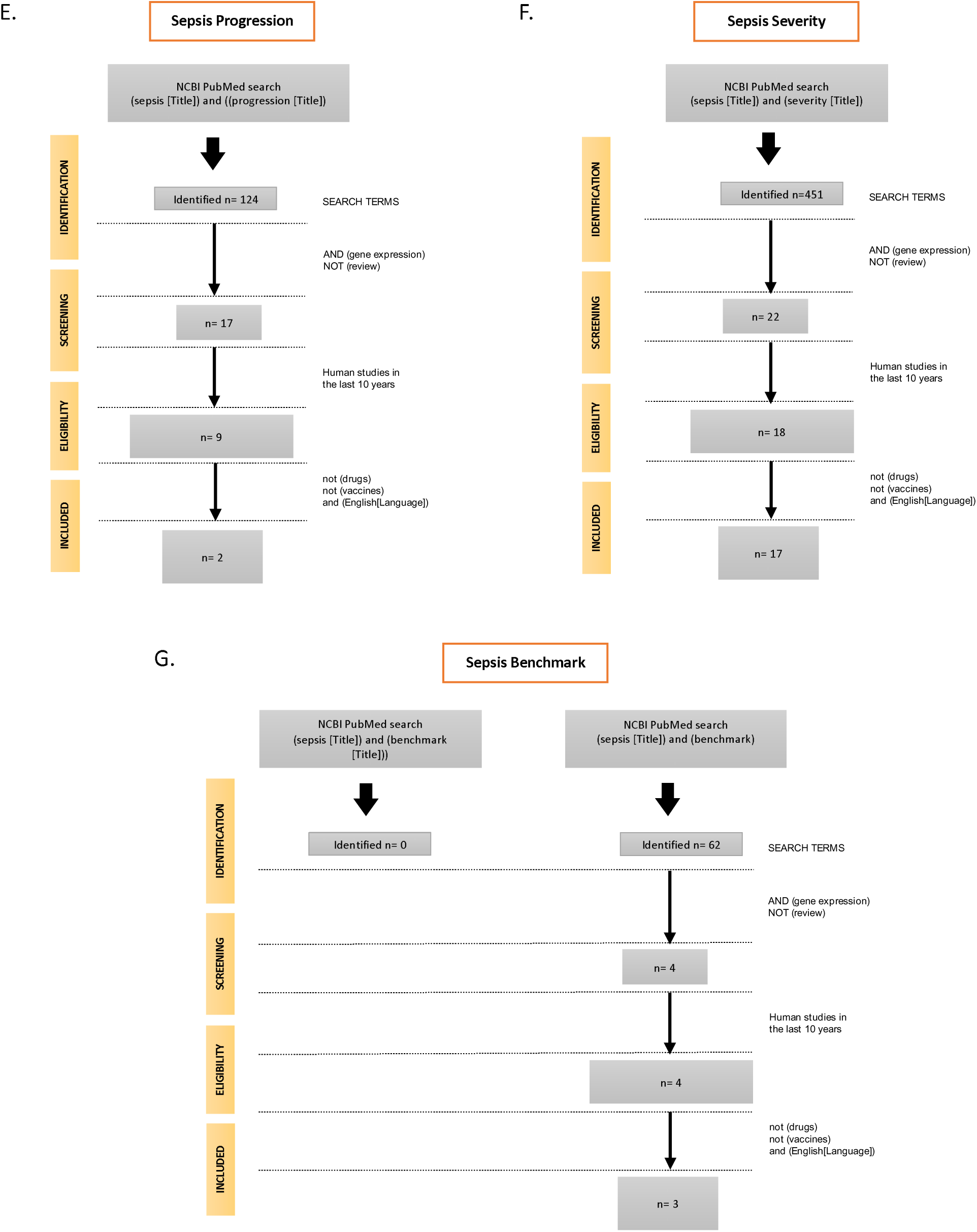
Search Terms. Studies were first selected (10^th^ July 2023) using the Pubmed web server according to selective keywords related to the framework headings (see methods). The identified studies were then screened, excluding review articles and including gene expression studies [Keyword Search Term: ‘(gene expression) not (review)’]. Human studies in the last ten years were then deemed eligible. Non-drug, vaccine, non-high throughput gene (HGT) studies and only research published in English were included in the final narrative analysis [Keyword Search term: not (drugs) not (vaccines) and (English)]. For the category **‘Sepsis Endotype’** the search strategy was changed as the title search only eluded to three studies; instead, a search through the text identified 64 studies, of which 17 were deemed eligible (2 were letters and comments to the editor and the third was a protein study). This left 14 included as HGT studies and for narrative review (Fig. 2A). In the category **‘Sepsis Biomarker’** 409 studies were identified after screening; this filtered the studies to 30, of which 23 were eligible, and six were included after exclusion (Fig. 2B). In the **‘Sepsis Definition’** category 125 studies were identified of which non were eligible after screening(Fig. 2C). For ‘**Sepsis Diagnosis’** 19 papers were deemed eligible, which after the exclusion, led to 15 studies of which only 7 were HGT studies (Fig. 2D). For **‘Sepsis Progression, ’** 124 studies were identified, 17 after screening, of which only nine were eligible, and two were included for narrative analysis (Fig. 2E). For ‘**Sepsis Severity**,’ 19 papers were eligible; after exclusion, this eluded 2 HGT studies (Fig. 2F). For ‘**Sepsis Benchmark’** the search strategy was changed to extend the search strategy through the body of the document (Fig. 2G).

**Figure 3:**
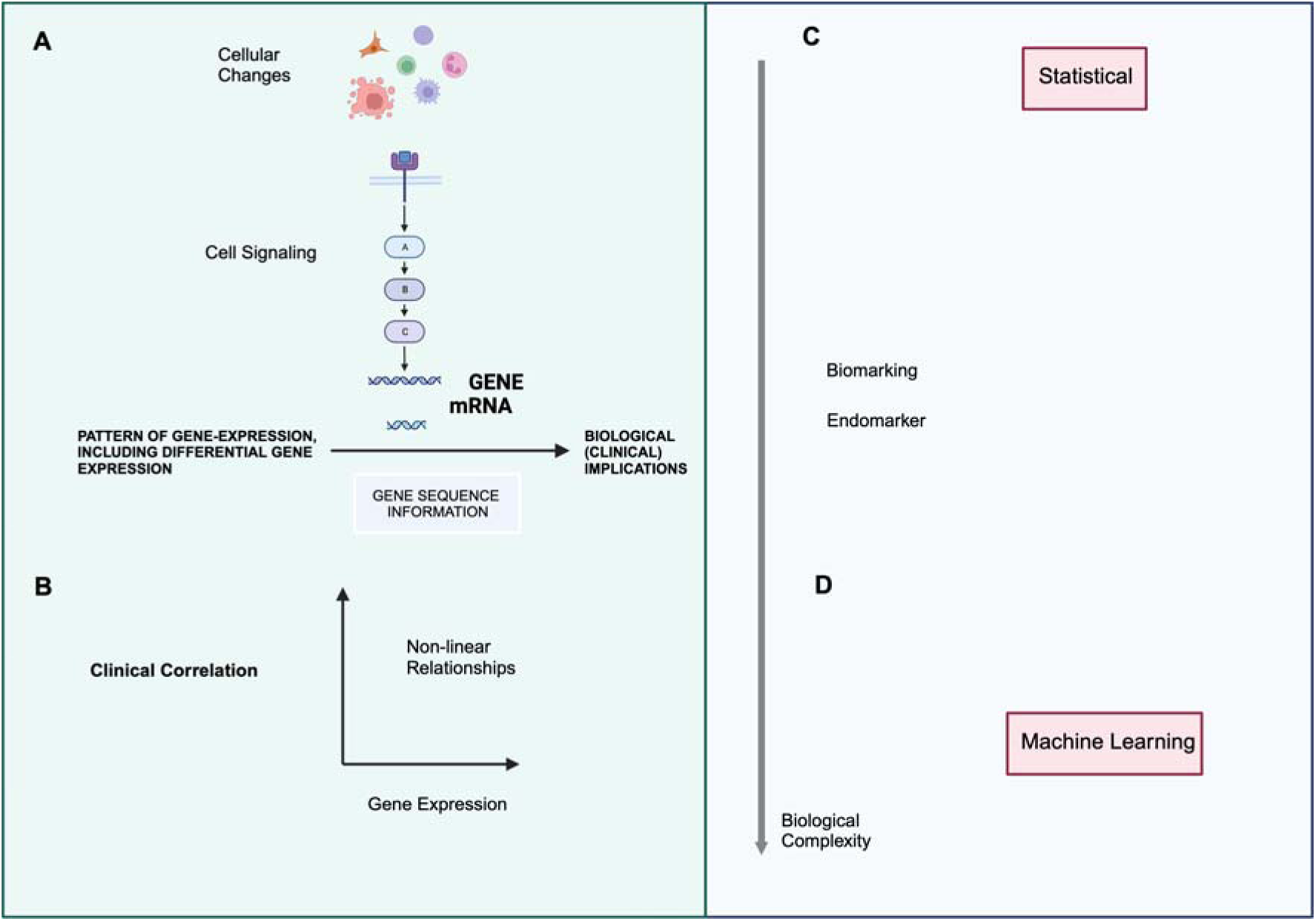
Correlation between changes at the cellular level, gene expression, and bioinformatic interpretation in sepsis. This figure explores the relationship between changes at the cellular level (left) to various data parameters (right) and their relationship to the disciplines of statistics for biological interpretation and Machine learning for data modeling is shown. **A.** Sepsis causes a multi-cellular response with associated activation of a complex immunological response. Various technologies are available to assess mRNA changes across the transcriptome (microarray, RNA-seq, sc-RNA-seq). This provides a holistic view of various processes and is advantageous over single-pathway approaches. **B.** However, given the heterogeneity of sepsis, the complex situation is challenging to analyze and develop from an omic perspective. Correlating clinical data to an omic perspective, such as gene expression, is complicated by the non-linear relationship of data points. **C.** Statistical method is required to adequately map biological changes at the cellular level, using gene expression data, to a bioinformatic interpretation. This has allowed the introduction of sepsis biomarkers. In an advancement from biomarking, the concept of endomarking allows the grouping of data according to functional biological characters. Statistical analysis requires a framework interpretation, which likely involves a degree of understanding of the data for simplification. **D.** Machine learning is useful as a modeling technique as it does not require a deep understanding of the inherent data structure. It is possible to apply various algorithms to the gene expression information and use various methods to train the data to develop a certain model. The model can then be tested with retrospective data and then validated prospectively.

### Study Inclusion and Exclusion Criteria

Articles were included in the review if they met three criteria: (1) focused on sepsis as the primary disease process and incorporated transcriptional (mRNA) analysis; (2) addressed one of five clinical areas of sepsis, including case definition, classification, Sepsis Severity Endotyping, Biomarkers and Benchmarking; and (3) involved human subjects.

Exclusions comprised conference abstracts and articles lacking full-text access or were not available in the English language. AR and JA independently screened the articles, resolving disagreements through discussion until consensus. Relevant information on applying transcript studies to sepsis was extracted and tabulated from the selected article using the Artificial Intelligence (AI) engine ‘Bing’ incorporated in the Microsoft Edge browser. The words “summarise the paper into header, methods, results and conclusions” were input into the AI engine, and the information generated was tabulated (Supplement: Tables 1 to 6). However, additional human scrutiny was applied to the tabulated studies with the discussion by the authors in the main text of the narrative review.

### Data Charting

We developed a data extraction form using Microsoft Excel (AR). Two independent reviewers (AR and JA) extracted data from the full-text articles to ensure consistency. The extracted data included population and study characteristics (e.g., demographics, aim, transcript information, significant genes, study outcomes, and conclusions).

### Data Collation and Result Reporting

The descriptive quantitative analysis undertaken in this study involved aggregating the number of articles based on key words in accordance to the aforementioned sepsis framework (Figure 1). The acquired studies were partitioned into two main framework categories. The first category encompassed concepts associated with cellular changes related to sepsis, such as endotypes and biomarkers. The second category addressed elements more directly related to the clinical presentation of sepsis, including its definition, diagnosis, progression, and severity. The results are subsequently presented according to this sequence, with a discussion of the relationship between the framework terms and gene expression.

## Results

The sepsis framework search terms identified 2,333 articles (Figure 2) and are detailed below (see headings) according to the framework search terms. A duplicate occurred once with the ‘Endotype’ and ‘Diagnosis’ searches and was recorded in both these sections.

### The Sepsis Endotype

"Endotype," derived from "endogenous phenotype," refers to disease subtypes defined by distinct biological mechanisms rather than just observable symptoms. Leveraging transcriptomic analysis can identify unique endotypes, transforming sepsis diagnosis and prognostics based on molecular characteristics and mechanisms.

Several studies have explored sepsis endotypes and mortality. Zhang et al. (2020) used deep learning to identify two sepsis classes - immunosuppressed Class 1 with higher mortality and relatively immunocompetent Class 2 with elevated mortality risk from hydrocortisone therapy ^26^. The VANISH trial found SRS2 endotype was associated with higher mortality when treated with corticosteroids ^27^. Pediatric sepsis had two endotypes (A and B), with Endotype A associated with higher 28-day mortality rates in patients with acute hypoxemic respiratory failure (AHRF) ^28^.

Transcriptomic analysis is valuable in understanding sepsis pathogenesis. Baghela et al. (2022) demonstrated gene expression signatures accurately predicting sepsis severity and identifying mechanistic endotypes in early sepsis ^29^. Got et al. (2020) linked sepsis-induced Epstein-Barr virus (EBV) reactivation to an immunosuppressed host transcriptomic endotype ^30^. Kwok et al. (2023) investigated neutrophils and emergency granulopoiesis in sepsis, finding altered gene expression in sepsis patients’ circulating hematopoietic stem and progenitor cells ^31^. Darden et al. (2021) used single-cell RNA sequencing to reveal the role of non-myeloid cells in chronic critical illness (CCI) and persistent inflammation, immunosuppression, and catabolism syndrome (PICS) after sepsis ^32^.

Combined with gene expression data, machine learning is a promising tool for identifying sepsis endotypes and improving prognosis. Sweeney et al. (2021) classified patients into Inflammopathic, Adaptive, and Coagulopathic endotypes, which are significantly associated with clinical outcomes and guiding personalized therapy ^33^. Banerjee et al. (2021) used Machine Learning to identify 20 differentially expressed genes predicting sepsis severity and outcome ^34^. Scicluna et al. (2017) identified four molecular endotypes (Mars1– 4) in sepsis patients, linked to severity scores, septic shock, and mortality ^35^.

Endotype-based research continues to evolve, supporting the potential for personalized sepsis management. Wong et al. (2012) developed ’PERSEVERE,’ a sepsis outcome prediction tool ^36^, while Lu et al. (2022) identified eight hub immune-related genes for sepsis diagnosis and prognosis ^37^. Baghela et al. (2023) utilized blood sepsis gene expression signatures to predict severity and endotypes in COVID-19 patients ^38^. They determined five endotypes reflecting distinct sepsis etiologies and therapeutic possibilities.

Efforts are underway to consolidate dysregulated gene sets linked to sepsis in the form of a library, such as ’SeptiSearch,’ a compendium of 103 unique gene sets developed by Baghela et al. (2023) ^39^. SeptiSearch includes a description of certain endotypes, thus included in this section. In summary, endotyping, along with machine learning techniques, holds significant promise for advancing sepsis management and delivering personalized care to patients.

### Sepsis Biomarker

Biomarkers, especially those derived from gene expression, are becoming increasingly vital in diagnosing and monitoring sepsis, providing measurable indicators of disease presence or severity. Among them, Zheng et al. (2020) distinguished bacterial and fungal sepsis via specific gene sets, introducing the bacterial sepsis Gene Set Variation Analysis (GSVA) index that exhibits remarkable discriminatory power between bacterial sepsis and non-sepsis samples ^40^. Moreover, Zhang et al. (2022) spotlighted the potential of ARG1 as a biomarker for diagnosing and prognosticating sepsis, linking ARG1 expression to disease severity and treatment response^41^. MicroaRNAs also demonstrate their utility in this field, with Huang et al. (2014) identifying eight novel miRNAs associated with the early diagnosis of sepsis ^42^, and Li et al. (2022) showcasing the diagnostic and prognostic value of BCL2A1 as a novel biomarker for sepsis management ^43^.

In a pivotal study, De Almeida et al. (2023) identified genes that linked Non-Thyroidal Illness Syndrome (NTIS) and sepsis, providing a critical insight into shared molecular mechanisms. Importantly, certain mitochondrial genes (mitGenes) stood out as potential survival prediction biomarkers. These mitGenes could differentiate between sepsis survivors and non-survivors, underlining the significant role they could play in sepsis endotyping. Among these, ROMO1, SLIRP, and TIMM8B emerged as potential predictive biomarkers of mortality in pediatric sepsis patients ^44^.

Transcriptomic biomarker panels also offer promise in the management of sepsis. Bauer et al. (2016) developed such a panel, which effectively quantified systemic inflammation and immune dysfunction in sepsis while also differentiating infected patients from those without infection. Additionally, this panel associated a down-regulated component of the genomic score with mortality ^45^.

### Sepsis Definition

The sepsis definition serves as a crucial basis for conveying research and clinical findings, with few studies taking on this challenge head-on. The Sepsis-3 committee acknowledged the complexity involved in matching the clinical physiological approach to the initial immunological changes in sepsis, noting that ambiguity in the definition could lead to inconsistent mortality reporting^17^.

The Sepsis-3 definition tackled this challenge by designating sepsis as a syndrome, recognizing the absence of a definitive diagnostic test. While the adult Sepsis-3 definition attempts to embody the intricacies of sepsis, it remains vague. It underscores the dysregulated immunological aspects of sepsis without fully being able to encompass the intricate and complex details. Transcriptomic perspectives could shed light on functional alterations in sepsis. For instance, Schaack et al. (2018) found distinct sepsis patient clusters exhibiting varying degrees of T-cell and monocyte functional loss, alongside dysregulated granulocytic neutrophil activation ^46^. Reyes et al. (2020), employing scRNA-seq analyses, identified 16 unique immune cell states, indicating that a transcriptomic functional interpretation of sepsis might aid in understanding its dysregulation. Nonetheless, a unifying immunological pattern that defines sepsis across various studies remains elusive^47^.

The pursuit of a comprehensive sepsis definition that encompasses age and pathogen type is a complex endeavor. Wynn et al. (2011) posited that age-related differences exist in septic shock, as neonates exhibit diminished gene expression in crucial immune-related pathways unlike other age groups^48^. This revelation prompts doubts about the viability of a universal, age-independent definition. Regarding the relationship of a sepsis definition according to pathogen type, research on SARS-CoV-2 has underscored the parallels between bacterial sepsis and COVID-19 dysregulated mechanisms. Karakike et al. (2021) reported that most ICU-hospitalized COVID-19 patients satisfied the Sepsis-3 criteria^49^.

Further, the emergence of SARS-CoV-2 has redirected research focus towards viral sepsis, unveiling shared features between bacterial sepsis and COVID-19 dysregulated mechanisms ^50^. Additionally, Sohn et al. (2020) suggested that the immune-related transcriptome profiles of COVID-19 patients mirrored those in bacterial sepsis, advocating for a pathogen-agnostic innate host response ^51^. Furthermore, Barh et al. (2020) showed that transcriptome studies of lung tissue post-SARS-CoV-2 infection revealed shared pathways with bacteria, parasites, and protozoa ^52^. These findings hint at the possibility of a pathogen-agnostic sepsis definition, even though its actualization remains riddled with hurdles ^50^. Innovative methods like transcriptomic approaches discussed in this review might be instrumental in bridging the gap in sepsis understanding, heading to an improved definition of sepsis.

### Sepsis Diagnosis

Kalantar et al. (2022) found that host gene expression from whole blood and plasma from 221 ICU patients could accurately differentiate sepsis from non-sepsis^53^. They used machine learning to develop classifiers based on host gene expression and pathogen detection. Combining host and microbial features improved sepsis diagnosis and predicted sepsis in patients with negative or indeterminate microbiological testing. Lukaszewski et al. (2022) identified specific gene signatures predicting infection or sepsis three days before clinical presentation^54^. Machine learning techniques accurately distinguished infection from uncomplicated recovery and sepsis from other postoperative presentations. Also, Xu et al. (2022) showed that microRNAs combined with TLR4/TDAG8 mRNAs and proinflammatory cytokines had utility as sepsis diagnosis biomarkers for early sepsis diagnosis^55^. Zhou et al. (2021) developed a 10-core gene expression panel for diagnosing pediatric sepsis, with ROC showing an AUC of the 10 core genes for diagnosing pediatric sepsis above 0.9 ^56^. Given the importance of the immune system in sepsis, Lu et al. (2022) focused on immune-related genes (IRGs) and their association with sepsis diagnosis and prognosis ^37^. Here, machine learning approaches identified hub IRGs from multiple datasets, establishing an IRG classifier based on 8 hub IRGs, which showed superior diagnostic efficacy and prognostic value compared to clinical characteristics alone (see the section on endotyping). The study also correlated the IRG classifier with immune-related characteristics, such as immune cell infiltration and cytokine expression. Sweeney et al (2018) validated a gene-expression test, the Sepsis Metacore (SMS), for sepsis in neonates. The SMS was accurate in three different cohorts of neonates with sepsis, and better than standard laboratory tests. Thereby suggesting that the SMS could help reduce unnecessary antibiotic use and improve outcomes for neonates with sepsis^57^.

### Sepsis Progression

The capacity to anticipate sepsis complications early, based on gene expression profiles, could offer the prospect of disease modification, allowing for individualized and targeted therapies. Fiorino et al (2022) undertook a prospective observational cohort study of 277 patients with infection, sepsis, or septic shock^58^. They used RNA sequencing of whole blood to measure the host gene expression response to infection and to identify signatures that could predict sepsis progression and mortality. The researchers found no gene expression signature for sepsis progression defined by the Sepsis-3 category, but they found signatures for sepsis progression defined by new organ dysfunction or ICU admission/mortality. They also validated four previously published gene signatures for sepsis mortality. By comparing the gene expression patterns of patients who progressed to more severe forms of sepsis or died within 28 days with those who did not, the authors identified genes and pathways associated with sepsis progression. Thus showing the ability to label sepsis progression based on host gene expression as a biomarker of the host response to infection. The elicited genes and molecular pathways could reflect different mechanisms (endotypes) of sepsis progression. The authors also used predictive modeling to generate gene expression signatures that classify patients into risk groups according to sepsis progression and mortality. Here a molecular score was provided according to g to the gene expression signature elicited, complementing clinical parameters to guide personalized approach to clinical care. Glibetic (2022) used transcriptomic analysis to identify patient subgroups with altered biological responses to sepsis, investigating the ethnic basis for viral infection risk and sepsis progression in colorectal cancer (CRC) patients^59^. Their analysis revealed distinct sepsis gene signatures classified as early and late response sepsis genes in the Native Hawaiian cohort compared to Japanese patients. Furthermore, canonical pathway analysis showed significant up and downregulation of mechanisms related to viral exit from host cells and epithelial junction remodeling. These findings suggest that genetic background plays a crucial role in sepsis heterogeneity, which could enable personalized approaches for risk stratification and targeted therapies.

### Sepsis Severity

De Jong et al. (2021) developed an innovative method to scrutinize disease-associated molecular changes using gene ensemble noise^60^. This measure, which represents the variance of gene groups, disrupts the conventional gene regulation model. The authors argue that cellular dynamics aren’t simply responsive to gene up- or down-regulation. Instead, they suggest the stochastic nature of gene expression impacts cellular responses. This approach allowed them to identify disturbances in sepsis-relevant pathways and protein complexes. They also noted its successful application to H1N1 infection and sepsis mortality. Their model predicted patient survival post-sepsis, and they incorporated WGCNA, emphasizing its value in understanding non-linear relationships. This approach predicted COVID-19 disease severity, revealing potential pharmaceutical targets.

In a parallel study, Baghela et al. (2022) sought gene expression signatures for sepsis severity and endotypes upon initial clinical presentation^29^. They proposed that sepsis is a syndrome comprising different endotypes, each representing a distinct group with unique severity and outcomes. Using whole blood RNA-Seq and machine learning, they analyzed gene expression profiles from ER and ICU patients with suspected sepsis. Their analysis identified five distinct endotypes with diverse underlying mechanisms. Two of them were associated with high severity and mortality, while one showed benign characteristics. The researchers developed a classification tool based on a multinomial regression model with LASSO regularization. This model, built on 40 genes, accurately predicted endotype status in sepsis patients and could be instrumental in early triage, potentially guiding personalized therapies. The study shed light on sepsis as a heterogeneous syndrome and provided valuable insights for predicting endotypes in sepsis patients.

### Sepsis Benchmark

Benchmarking, the process of comparing a system’s or method’s performance against a recognized standard, plays a crucial role in clinical research, especially in sepsis studies ^61^. However, the core challenge is identifying the correct standard against which to benchmark. One solution involves classifying gene patterns based on disease conditions or processes, thereby creating a reference library of gene patterns. In this vein, Altman et al. (2021) developed a transcriptomic benchmarking framework called BloodGen3Module, designed to facilitate the analysis of gene expression data^62^.

In a notable study by Sweeney et al. (2017), three gene expression diagnostic classifiers, namely the 11-gene Sepsis MetaScore, FAIM3:PLAC8 ratio, and the Septicyte Lab, were tested on 39 publicly available sepsis datasets ^63^. The objective was to determine how well these classifiers could distinguish patients with infection from those with non-infectious inflammation. The three diagnostic classifiers performed similarly in separating non-infectious SIRS from sepsis, but the Septicyte Lab performed less well in separating infections from healthy controls. In a subsequent study, Sweeney et al. (2018) conducted a validation study of the Sepsis MetaScore for diagnosing sepsis in neonates, demonstrating its superior performance over standard laboratory measurements across three distinct cohorts ^57^.

Another significant contribution to sepsis benchmarking was made by Scicluna et al. (2020). The authors carried out a next-generation microarray analysis of leukocyte RNA from 156 patients with sepsis and 82 healthy subjects, eight of whom underwent a lipopolysaccharide challenge in a clinically controlled setting, a process known as human endotoxemia ^64^. The study found significant alterations in long non-coding RNA and, to a lesser extent, small non-coding RNA, in sepsis patients compared to healthy subjects. Moreover, their results highlighted the potential relevance of non-sensory olfactory receptor activity among other pathways, and suggested that long non-coding RNA profiles in sepsis could serve as a benchmark for future studies.

While sepsis benchmarking remains invaluable, it is not without challenges. The need for well-labeled transcriptomic data and the necessity of a universally accepted definition of sepsis across studies are significant hurdles. Nonetheless, sepsis benchmarking continues to be a vital tool in the evaluation of gene transcriptomics tools and methodologies, serving to guide researchers in their quest for effective prediction methods, biomarker discovery, and the development of novel therapeutics.

## Discussion

A system-wide approach, using transcriptomic analysis, was undertaken connecting conventional sepsis themes as a part of ten-year literature scoping review. Hence, a transcriptomics-oriented approach was proposed using gene expression studies spanning various categories—diagnostic, organ dysfunction, sepsis severity, endotyping, classification, biomarking, and benchmarking. In the presented framework, the terms were broadly grouped into two categories. The first provides scientific insights based on changes at the molecular or cellular level (Biomarkers and Endotypes). The other category provided more high-level insights appropriate to a clinical understanding. The interplay between scientific (biological) or clinical terms to features of genomic data seemed intuitive.

Biomarkers and endotypes formed components of the scientific category. A biomarker refers to objectively measurable signs, such as molecules found in the blood or changes in body function, that indicate biological or pathogenic processes or responses to therapeutic intervention ^65^. Conversely, an endotype is employed in classification, highlighting the connection between a disease and a distinct pathophysiological mechanism^66^. Biomarkers have proven their worth in sepsis studies, providing a convenient and objective method for disease observation, tracking, and patient response monitoring^67^. Nonetheless, endotypes could offer a more profound bridge between the biological and clinical contexts. Endotypes, representing distinct underlying disease pathways and mechanisms, are typically identified using advanced diagnostic techniques like genetic analysis, molecular profiling, and biomarker identification. By discerning specific gene expression patterns or molecular signatures unique to a patient subgroup within a broader disease category, endotypes enable more personalized treatment strategies. Continuous research and technological advancements promise to refine our understanding of endotypes, heralding a future of increasingly personalized and effective medical care.

The disparity between scientific insights in sepsis and their clinical application remains a limitation across sepsis studies in general. Factors contributing to the difficulty in transferring insights across RNA-based studies, including dealing with the variability due to platform heterogeneity, sample collection timing, and host-pathogen differences, including demographic aspects. These factors make the linkage of different studies of sepsis problematic. Most importantly, a clear definition of sepsis is a major obstacle in creating an effective research framework. This may be the reason for a further limitation of this study due to the blurred boundaries between research terms, especially those related to clinical descriptors such as sepsis severity and progression. Moreover, although there is abundant research on the use of the chosen framework terms, the lack of a universally accepted structure to ensure their clinical relevance added to the complexity.

Future development should bridge the two domains, scientific and clinical, in forwarding sepsis research. This would facilitate the application of omic approaches to clinical practice. Then this could allow a more seamless application of omics-based methods, such as transcriptomics, to fill knowledge gaps. Hence the application of high-resolution metrics based on gene expression technologies for personalized diagnostics in sepsis should remain a future research goal. Moreover, given the likely highly non-linearity of gene expression data, future research could explore the application of artificial intelligence to further address the issue of ambiguity. Advanced algorithms and machine learning techniques could help interpret complex gene expression data and other omics information, offering further clarity and precision to disease classification and treatment strategies. These strategies may be instrumental in overcoming the inherent challenges and moving closer to more precise and personalized care for sepsis patients. Advancements in defining sepsis and identifying biomarkers can positively impact benchmarking, thereby extending applicability across various clinical sepsis realms. Moreover, the exciting development of technologies towards faster processing of high throughput gene expression data may allow the application of transcriptomics in acute sepsis. Importantly, gene expression methods, as detailed in this study, hold the future promise of validating a consensus-driven approach for sepsis management, bridging existing knowledge gaps in sepsis pathophysiology and allowing the refinement of clinical protocols using precision strategies.

## Conclusion

This narrative review underscored the potential of a transcriptomics-oriented approach as a pivotal tool for bridging knowledge gaps between pathophysiological changes, and cellular modifications applied to the clinical context. The transcriptomics-oriented approach used gene expression studies spanning various sepsis categories—diagnostic, progress, severity, endotyping, classification, biomarking, and benchmarking. This revealed the application of transcriptomics across numerous aspects of sepsis, offering promising avenues for integration with other omic strategies and interpretive frameworks. Nonetheless, the inherent complexities and interpretive challenges of sepsis persistently echoed throughout the review. Future omic research should look at sealing the gaps between biological changes translated to the clinical context. By adopting a transcriptomic-driven approach, researchers and clinicians can collectively navigate the intricacies of sepsis, directing future progress and facilitating improved patient outcomes from sepsis.

## Author Contributions

Conceived and designed the methodology: AR

Performed the experiments: AR

Analyzed the data: AR, PAC

Contributed analysis, methods, and tools: AR

Wrote the first draft of the paper: AR

Supervision: MGS, AA, AH

Revised critically for importance and intellectual content: AR, KW, HA, BH, PK, AS,CK, BSB, MT, ZH, HV, GB, ZM, RN, RM, SD, NK, RK, AS, PW, MGS, JS, SAZ, RP, MT, WZ, MAZ, HS, AA, AH

## Data Availability

All cited literature has been used for this scoping review and is available as per the references provided according to journals in the public domain.

## Acknowledgments

The authors thank the anonymous reviewers for their insightful comments and suggestions. Nuha Kidwai, North London Collegiate, Dubai for online search term testing. Hussain acknowledges the support of the UK Engineering and Physical Sciences Research Council (EPSRC) - Grants Ref. EP/M026981/1, EP/T021063/1, EP/T024917/1. For Dr. Binu George for keeping us on track with the study goals. To Professor Hector Wong, a pioneer in the field of transcriptomics of sepsis; to his enduring contribution to the field.

---

**Supplement Table 1.** Endotype. Details the search results according to the chosen sepsis framework search term ‘Endotype’. * Literature Reference ** This paper appears in ‘Endotype’ and ‘Diagnosis’ searches.

| Study | Description | Study Conclusion | Ref* |
| --- | --- | --- | --- |
| Deep learning-based clustering robustly identified two classes of sepsis with both prognostic and predictive values | The study aimed to develop a parsimonious class model to predict class membership and validate the model for its prognostic and predictive capability in external datasets. The Gene Expression Omnibus (GEO) and ArrayExpress databases were searched from inception to April 2020. Datasets containing whole blood gene expression profiling in adult sepsis patients were included. Autoencoder was used to extract representative features for k-means clustering. Genetic algorithms (GA) were employed to derive a parsimonious 5-gene class prediction model. The class model was then applied to external datasets (n = 780) to evaluate its prognostic and predictive performance. The study found that 12 datasets involving 1613 patients were included. Two classes were identified in the discovery cohort (n = 685). Class 1 was characterized by immunosuppression with higher mortality than class 2 (21.8% [70/321] vs. 12.1% [44/364]; $p < 0.01$ for Chi-square test). A 5-gene class model (C14orf159, AKNA, PILRA, STOM, and USP4) was developed with GA. In external validation cohorts, the 5-gene class model (AUC: 0.707; 95% CI: 0.664 – 0.750) performed better in predicting mortality than sepsis response signature (SRS) endotypes (AUC: 0.610; 95% CI: 0.521 – 0.700) and performed equivalently to the APACHE II score (AUC: 0.681; 95% CI: 0.595 – 0.767). In the dataset E-MTAB-7581, using hydrocortisone was associated with an increased mortality risk (OR: 3.15 [1.13, 8.82]; $p = 0.029$) in class 2. | The study concludes that their study identified two classes of sepsis that showed different mortality rates and responses to hydrocortisone therapy. Class 1 was characterized by immunosuppression with a higher mortality rate than class 2. They further developed a 5-gene class model to predict class membership. | 26 |
| Predicting sepsis severity at first clinical presentation: The role of endotypes and mechanistic signatures | The study aimed to identify gene expression signatures that predict subsequent severity in sepsis patients at first clinical presentation. Blood RNA-Seq and clinical data were collected from 348 patients in four emergency rooms (ER), one intensive-care unit (ICU), and 44 healthy controls. Gene expression profiles were analyzed using machine learning and data mining to identify clinically relevant gene signatures reflecting disease severity, organ dysfunction, mortality, and specific endotypes/mechanisms. The study found that gene expression signatures were obtained that predicted severity/organ dysfunction and mortality in both ER and ICU patients with an accuracy/AUC of 77-80%. Network analysis revealed these signatures formed a coherent biological program with specific but overlapping mechanisms/pathways. Patients with early sepsis could be stratified into five distinct and novel mechanistic endotypes, named Neutrophilic-Suppressive/NPS, Inflammatory/INF, Innate-Host-Defense/IHD, Interferon/IFN, and Adaptive/ADA, each based on approximately 200 unique gene expression differences, and distinct pathways/mechanisms. Endotypes had varying overall severity, with two severe (NPS/INF) and one relatively benign (ADA) groupings. A 40-gene classification tool (accuracy=96%) and several gene pairs (accuracy=89-97%) accurately predicted endotype status in both ER and ICU validation cohorts. | The study concludes that the severity and endotype signatures indicate that distinct immune signatures precede the onset of severe sepsis and lethality, providing a method to triage early sepsis patients. | 29 |
| Classification of patients with sepsis according to blood genomic endotype: a prospective cohort study | This prospective observational cohort study included consecutive patients admitted for sepsis to two intensive care units (ICUs) in the Netherlands and patients admitted with sepsis due to community-acquired pneumonia to 29 ICUs in the UK. The researchers generated genome-wide blood gene expression profiles from admission samples and analyzed them using unsupervised consensus clustering and machine learning. Four molecular endotypes for sepsis designated Mars1–4, were identified in the discovery cohort and were associated with 28-day mortality. The discovery cohort found the worst outcome for patients classified as having a Mars1 endotype. A 140-gene expression signature reliably stratified patients with sepsis to the four endotypes in the first and second validation cohorts. Only Mars1 was consistently significantly associated with 28-day mortality across the cohorts. | This study provides a method for the molecular classification of patients with sepsis to four different endotypes upon ICU admission. Detection of sepsis endotypes might assist in personalized patient management and selection for trials. | 35 |
| A Novel Single Cell RNA-seq Analysis of Non-Myeloid Circulating Cells in Late Sepsis | This study used single-cell RNA sequencing (scRNA-seq) to perform a detailed transcriptomic analysis of lymphoid-derived leukocytes to better understand the pathology of late sepsis. A mixture of whole blood myeloid-enriched and Ficoll-enriched peripheral blood mononuclear cells from four late septic patients (post-sepsis day 14-21) and five healthy subjects underwent Cellular Indexing of Transcriptomes and Epitopes by Sequencing (CITE-seq). The study identified unique transcriptomic patterns for multiple circulating immune cell subtypes, including B- and CD4+, CD8+, activated CD4+ and activated CD8+ T-lymphocytes, as well as natural killer (NK), NKT, and plasmacytoid dendritic cells in late sepsis patients. The analysis demonstrated that the circulating lymphoid cells maintained a transcriptome reflecting immunosuppression and low-grade inflammation. The study also identified transcriptomic differences between patients with bacterial versus fungal sepsis, such as greater expression of cytotoxic genes among CD8+ T-lymphocytes in late bacterial sepsis. | Circulating non-myeloid cells display a unique transcriptomic pattern late after sepsis. Non-myeloid leukocytes in particular, reveal a host endotype of inflammation, immunosuppression, and dysfunction, suggesting a role for precision medicine-guided immunomodulatory therapy | 32 |
| Transcriptomic Signatures in Sepsis and a Differential Response to Steroids From the VANISH Randomized Trial | This was a post hoc analysis of a double-blind, randomized clinical trial in septic shock (VANISH [Vasopressin vs. Norepinephrine as Initial Therapy in Septic Shock]). Patients were included within 6 hours of the onset of shock and were randomized to receive norepinephrine or vasopressin followed by hydrocortisone or placebo. Genome-wide gene expression profiling was performed, and the SRS endotype was determined by a previously established model using seven discriminant genes. Samples were available from 176 patients: 83 SRS1 and 93 SRS2. There was no significant interaction between the SRS group and vasopressor assignment ($P = 0.50$). However, there was an interaction between assignment to hydrocortisone or placebo and SRS endotype ($P = 0.02$). Hydrocortisone use was associated with increased mortality in those with an SRS2 phenotype (odds ratio = 7.9; 95% confidence interval = 1.6–39.9). | Transcriptomic profile at onset of septic shock was associated with response to corticosteroids. Those with the immunocompetent SRS2 endotype had significantly higher mortality when given corticosteroids than placebo. | 27 |
| Epstein-Barr virus reactivation in sepsis due to community-acquired pneumonia is associated with increased morbidity and an immunosuppressed host transcriptomic endotype | This study aimed to determine the incidence and correlates of Epstein-Barr virus (EBV) positivity in a large sepsis cohort. The researchers assayed EBV from the plasma of intensive care unit (ICU) patients with sepsis due to community-acquired pneumonia. In total, 730 patients were evaluated by targeted metagenomics, digital droplet PCR, or both. Gene expression in peripheral blood leukocytes was analyzed for a subset of individuals.<br>The study found a 37% incidence of EBV-positivity. EBV reactivation was associated with longer ICU stay and increased organ failure. EBV reactivation was also associated with the relatively immunosuppressed SRS1 endotype and differential expression of a few biologically relevant genes. | These findings are consistent with the hypothesis that viral reactivation in sepsis results from immune compromise and is associated with increasing severity of illness. However, further mechanistic studies are required to definitively illustrate cause and effect. | 30 |
| EVIDENCE OF ENDOTYPES IN PEDIATRIC ACUTE HYPOXEMIC RESPIRATORY FAILURE CAUSED BY SEPSIS | This study was a secondary analysis of a previously reported microarray-based study of pediatric sepsis. The researchers tested whether endotypes identified in pediatric sepsis apply to acute hypoxemic respiratory failure (AHRF). The study was conducted in multiple pediatric intensive care units in the United States, including 67 children with AHRF caused by sepsis.<br>Of the larger septic shock cohort, 67 met eligibility for AHRF. Twenty-three subjects were assigned to Endotype A, and 44 to Endotype B. Subjects assigned to Endotype A had over 4-fold greater unadjusted 28-day mortality and nearly 3-fold greater rates of complicated course. The association with mortality and complicated course persisted after adjustment for age, severity of illness, and PaO<sub>2</sub>/FIO<sub>2</sub>. | Applying a previously reported endotyping strategy in children with septic shock identified endotypes of pediatric AHRF secondary to sepsis, with differential risk for poor outcomes. This is the first demonstration of endotypes in pediatric respiratory failure. The results support an investigation into using transcriptomics to identify mRNA-based endotypes in a dedicated, well-defined AHRF cohort. | 28 |
| Facilitating systems-level analyses of all-cause and Covid-mediated sepsis through SeptiSearch, a manually-curated compendium of | This study aimed to build a compendium of previously described gene sets that combine knowledge from sepsis-associated studies. PubMed was searched for transcriptomics studies to characterize acute infection/sepsis and severe sepsis. The molecules included in each gene set were collected in addition to the relevant study metadata.<br>After extensive literature curation of 74 sepsis-related publications involving transcriptomics, 103 unique gene sets (comprising | SeptiSearch provides members of the sepsis community with the bioinformatic tools needed to leverage and explore the gene sets contained in the database. This will allow the gene sets to be further scrutinized and analyzed for | 39 |
| dysregulated gene sets | 20,899 unique genes) from thousands of patients were collated with associated metadata. Frequently described genes included in gene sets and the molecular mechanisms they were involved in were identified. The SeptiSearch database is available in a web application created using the Shiny framework in R. | their enrichment in user-submitted gene expression data and used to validate in-house gene sets/signatures. |  |
| Machine Learning Identifies Complicated Sepsis Course and Subsequent Mortality Based on 20 Genes in Peripheral Blood Immune Cells at 24 H Post-ICU Admission | The authors identified 20 genes differentially expressed between complicated and uncomplicated sepsis patients and developed a highly accurate machine-learning model that could predict complicated course outcomes and mortality. They also found eight novel (CLEC5A, TCN1, HCAR3, MS4A3, OLAH, PLCB1, SDC4, and NLRP1) genes associated with sepsis's overactive innate immune system and neutrophil function.<br>In this study, the artificial intelligence model used was a combination of variable selection techniques, oversampling and undersampling methods, and tree-based classifiers and logistic regression. The authors used three variable selection techniques (random forest-based variable importance, LASSO, and Minimum Redundancy and Maximum Relevance) to reduce the dimensionality of the gene expression data and identify the most important genes. They also used different oversampling and undersampling techniques (such as SMOTE, Cluster Centroids, and Instance Hardness Threshold) to balance the imbalanced data<sup>1</sup>[1]. They then developed machine learning algorithms using tree-based classifiers (such as Balanced Random Forest and Extra Trees) and logistic regression to classify the patients into complicated or uncomplicated course outcomes. They tuned the hyperparameters of the classifiers using a cross-validated grid search technique and evaluated their performance using metrics such as AUROC, sensitivity, specificity, and Mathews correlation coefficient. They also experimented with different classification thresholds to fine-tune the classifiers for imbalanced data. They repeated the process 10 times with 5-fold cross-validation to generate 50 iterations and averaged the results. They also validated their model and genes using similar methods on four external datasets. | The researchers proposed a new method and biomarkers for early identification of sepsis disease trajectory and clinical outcomes, which could help modify treatment options and improve patient survival. | 34 |
| Pediatric Sepsis Endotypes among Adults with Sepsis | The authors used transcriptomic data from adults with sepsis from the GAINs study and assigned them to SRS groups and endotype groups using previously reported gene expression signatures. They used multivariable logistic regression to assess the associations between endotype, SRS membership, age, and mortality.<br>The researchers found that 30% of the subjects were assigned to endotype A and 70% to endotype B. A weak, positive correlation existed between endotype assignment and SRS membership. SRS1 membership, endotype A assignment, and older age were associated with an increased risk of mortality. The interaction between endotype and age was statistically significant, showing an increased mortality risk with endotype A and younger age. Subjects co-assigned to endotype A and SRS1 had the highest mortality. Pathway analysis revealed that the genes differentiating the A1 group from the others corresponded to the T cell receptor signaling pathway. | The authors concluded that the SRS and endotyping strategies might provide complementary, age-dependent biological and prognostic information for sepsis sub-classification. They suggested future studies directly assessing the specific biology reflected by these two signatures and how they interact in sepsis pathobiology. | 69 |
| Characterization of immune-related genes and immune infiltration features for early diagnosis, prognosis and recognition of immunosuppression in sepsis** | This study used a combination of machine learning approaches, including modified Lasso penalized regression and random forest (RF), to identify hub immune-related genes (IRGs) from multiple global datasets. The IRG score was generated through a linear combination of coefficients from logistic regression and the relative expression of each IRG. The diagnostic and prognostic values of the model were tested in independent validation cohorts from ArrayExpress databases. The authors also systematically correlated the IRG classifier with immunological characteristics from multiple perspectives, such as immune-related cells infiltrating, pivotal molecular pathways, and cytokine expression.<br>The study identified 8 hub IRGs (ADM, CX3CR1, DEFA4, HLA- | The study established a diagnostic and prognostic model based on 8 IRGs that is closely correlated with responses to hydrocortisone and immunosuppression status. This model might facilitate personalized counseling for specific therapy. The results suggest that components of the immune system play a crucial role in the occurrence and development of sepsis. | 37 |
|  | DPA1, MAPK14, ORM1, RETN, and SLPI) that were combined to construct an IRG classifier. In the discovery cohort, the IRG classifier exhibited superior diagnostic efficacy and performed better in predicting mortality than clinical characteristics or MARS/SRS endotypes. Encouragingly, similar results were observed in the ArrayExpress databases. The use of hydrocortisone in the IRG high-risk subgroup was associated with increased risk of mortality. In the IRG low-risk phenotypes, NK cells, T helper cells, and infiltrating lymphocyte (IL) were significantly richer, while T cells regulatory (Tregs) and myeloid-derived suppressor cells (MDSC) were more abundant in the IRG high-risk phenotypes. |  |  |
| Predicting severity in COVID-19 disease using sepsis blood gene expression signatures | The authors analyzed the whole blood transcriptome of 124 patients with confirmed COVID-19 infections from hospitals in Quebec, Canada<sup>[1]</sup>. They compared the gene expression profiles of patients with different severity levels and outcomes, and assessed the dysregulation of sepsis signatures and endotypes that reflect distinct pathophysiology and underlying mechanisms.<br>The authors found that signatures reflecting cellular reprogramming, organ dysfunction, and mortality were significantly enriched and predictive of severity and lethality in COVID-19 patients. They also identified five endotypes that reflected distinct sepsis aetiologies and therapeutic opportunities. The endotypes were associated with different clinical parameters, such as respiratory support group, ICU admission, and mortality. The authors also identified potential endotype-specific drug targets using protein-protein interaction networks. | The authors suggested that severe COVID-19 patients should be classified as having severe sepsis and that sepsis signatures could be useful for clinical risk stratification and treatment guidance. They also proposed that endotypes might enable personalized medicine approaches for COVID-19 disease. | 38 |
| Validation of Inflammopathic, Adaptive, and Coagulopathic Sepsis Endotypes in Coronavirus Disease 2019 | The authors applied a 33-messenger RNA classifier to assign endotype (Inflammopathic, Adaptive, or Coagulopathic) to 97 patients with COVID-19 within 24 hours of hospital admission<sup>[1]</sup>. They tested endotype status against other clinical parameters including laboratory values, severity scores, and outcomes.<br>The authors found that endotype assignment was associated with clinical outcomes and immune markers. They showed that the Adaptive endotype had no deaths, while the Inflammopathic and Coagulopathic endotypes had high mortality rates<sup>[3]</sup>. Age and endotype probability were significant predictors of death in a multivariate regression model. The authors also explored the relationship between endotypes and coagulation parameters, such as d-dimers, fibrinogen, and ISTH DIC score. They observed significant differences across the endotypes, with the Coagulopathic group showing the highest levels of coagulopathy markers. | The authors suggest that the heterogeneity of sepsis is also present in COVID-19. The 33-messenger RNA classifier is one possible way to reduce the clinical heterogeneity and potentially inform therapeutic decisions. They suggest that future studies of immunomodulatory therapy in sepsis or COVID-19 should consider the endotypes as a companion-diagnostic approach. | 33 |
| Neutrophils and emergency granulopoiesis drive immune suppression and an extreme response endotype during sepsis | The authors used single-cell multiomic analysis to profile the blood cells of patients with sepsis and healthy controls, and identified different subsets of neutrophils and hematopoietic stem and progenitor cells (HSPCs) that were altered in sepsis<sup>[2]</sup>. They also performed functional assays to test the immunosuppressive properties of neutrophils and the granulopoietic profiles of HSPCs. They further analyzed the relationship between the neutrophil and HSPC features and the sepsis response signatures (SRSs), which are transcriptomic subphenotypes of sepsis associated with different outcomes and responses to infection.<br>They found that neutrophils and emergency granulopoiesis, the rapid production of granulocytes from HSPCs, drove a maladaptive response during sepsis that resulted in immune suppression and an extreme response endotype, a subgroup of patients with poor outcome and a specific sepsis response signature (SRS1). They showed that neutrophils from patients with sepsis inhibited the proliferation and activation of T cells, and that this effect was mediated by prostaglandins and genes associated with granulocytic myeloid-derived suppressor cells (G-MDSCs). They also showed that circulating HSPCs from patients with sepsis had altered granulopoietic profiles, with increased expression of genes and transcription factors involved in emergency granulopoiesis and expanded myelopoiesis. | They concluded that their findings revealed potential therapeutic targets and opportunities for stratified medicine in severe infection. They suggested that modulating neutrophil function or granulopoiesis could improve the outcome of patients with sepsis, especially those with the SRS1 subphenotype. | 31 |

**Table 2.** Biomarker. Details the search results according to the chosen sepsis framework search term ‘Biomarker’. * Literature Reference

| Study | Descriptions | Study Conclusion | Ref* |
| --- | --- | --- | --- |
| Two Gene Set Variation Index as Biomarker of Bacterial and Fungal Sepsis | The study applied microarray data of bacterial sepsis, fungal sepsis, and mock-treated samples to perform differentially expressed gene (DEG) analysis to identify a bacterial sepsis-specific gene set and a fungal sepsis-specific gene set. Functional enrichment analysis was used to explore the body's response to bacterial sepsis and fungal sepsis. Gene set variation analysis (GSVA) was used to score individual samples against the two pathogen-specific gene sets, and each sample gets a GSVA index. Receiver operating characteristic (ROC) curve analysis was performed to evaluate the diagnostic value of sepsis. An independent data set was used to validate the bacterial sepsis-specific GSVA index.<br>The study found that the genes differentially expressed only in bacterial sepsis and the genes differentially expressed only in fungal sepsis were significantly involved in different biological processes (BPs) and pathways. This indicated that the body's responses to fungal sepsis and bacterial sepsis are varied. Twenty-two genes were identified as bacterial sepsis-specific genes and upregulated in bacterial sepsis, and 23 genes were identified as fungal sepsis-specific genes and upregulated in fungal sepsis. ROC curve analysis showed that both of the two pathogen sepsis-specific GSVA indexes may be a reliable biomarker for corresponding pathogen-induced sepsis (AUC = 1:000), while the mRNA of CALCA (also known as PCT) have a poor diagnostic value with AUC = 0:512 in bacterial sepsis and AUC = 0:705 in fungi sepsis. In addition, the AUC of the bacterial sepsis-specific GSVA index in the independent data set was 0.762. | The study concludes that they proposed a bacterial sepsis-specific gene set and a fungal sepsis-specific gene set; the bacterial sepsis GSVA index may be a reliable biomarker for bacterial sepsis. | 40 |
| ARG1 as a promising biomarker for sepsis diagnosis and prognosis: evidence from WGCNA and PPI network | The study aimed to identify crucial genes and biomarkers for sepsis that could guide clinicians to make rapid diagnosis and prognostication. Preliminary analysis of multiple global datasets, including 170 samples from patients with sepsis and 110 healthy control samples, revealed common differentially expressed genes (DEGs) in peripheral blood of patients with sepsis. After Gene Ontology (GO) and pathway analysis, the Weighted Gene Correlation Network Analysis (WGCNA) was used to screen for genes most related with clinical diagnosis. Also, the Protein-Protein Interaction Network (PPI Network) was constructed based on the DEGs and the hub genes were found. The results of WGCNA and PPI network were compared and one shared gene was discovered. Then more datasets of 728 experimental samples and 355 control samples were used to prove the diagnostic and prognostic value of this gene. Last, real-time PCR was used to confirm the bioinformatic results.<br>The study found that four hundred forty-four common differentially expressed genes in the blood of sepsis patients from different ethnicities were identified. Fifteen genes most related with clinical diagnosis were found by WGCNA, and 24 hub genes with most node degrees were identified by PPI network. ARG1 turned out to be the unique overlapped gene. Further analysis using more datasets showed that ARG1 was not only sharply up-regulated in sepsis than in healthy controls, but also significantly high-expressed in septic shock than in non-septic shock, significantly high-expressed in severe or lethal sepsis than in uncomplicated sepsis, and significantly high-expressed in non-responders than in responders upon early treatment. These all demonstrate the performance of ARG1 as a key biomarker. Last, the up-regulation of ARG1 in the blood was confirmed experimentally. | The study concludes that they identified crucial genes that may play significant roles in sepsis by WGCNA and PPI network. ARG1 was the only overlapped gene in both results and could be used to make an accurate diagnosis, discriminate the severity and predict the treatment response of sepsis. | 41 |
| Similar hypothyroid and sepsis circulating mRNA expression could be useful as a biomarker in nonthyroidal illness syndrome: a pilot study | The study aimed to compare the patterns of gene expression between hypothyroidism and nonthyroidal illness syndrome (NTIS) as models of NTIS. The researchers used Ion Proton System next-generation sequencing to build the hypothyroidism transcriptome. They selected two databanks in GEO2 platform datasets to find the differentially expressed genes (DEGs) in adults and children with sepsis. The ROC curve was constructed to calculate the area under the curve (AUC). The AUC, chi-square, sensitivity, specificity, accuracy, kappa and likelihood were calculated. Cox regression and Kaplan-Meier analyses were performed for the survival analysis.<br>The study found that concerning hypothyroidism DEGs, 70.42% were shared with sepsis survivors and 61.94% with sepsis nonsurvivors. Some of them were mitochondrial gene types (mitGenes), and 95 and 88 were related to sepsis survivors and nonsurvivors, respectively. BLOC1S1, ROMO1, SLIRP and TIMM8B mitGenes showed the capability to distinguish sepsis survivors and nonsurvivors. | The study concludes that they matched their hypothyroidism DEGs with those in adults and children with sepsis. Additionally, they observed different patterns of hypothyroid-related genes among sepsis survivors and nonsurvivors. Finally, they demonstrated that ROMO1, SLIRP and TIMM8B could be predictive biomarkers in children's sepsis. | 44 |
| Identification of MicroRNA as Sepsis Biomarker Based on miRNAs Regulatory Network Analysis | The study aimed to identify novel microRNA biomarkers associated with the early diagnosis of sepsis by analyzing miRNA expression profiles and the miRNA regulatory network. Pathways analysis, disease ontology analysis, and protein-protein interaction network (PIN) analysis, as well as ROC curve, were exploited to testify the reliability of the predicted miRNAs.<br>The study found that by analyzing the miRNA expression profiles and the miRNA regulatory network, they obtained novel miRNAs associated with sepsis. They finally identified 8 novel miRNAs which have the potential to be sepsis biomarkers. | The study concludes that by applying a miRNA regulatory network-based method, they were able to identify novel microRNA biomarkers associated with the early diagnosis of sepsis. These findings may have important implications for improving the diagnosis and treatment of sepsis. | 42 |
| Bulk RNA Sequencing With Integrated Single-Cell RNA Sequencing Identifies BCL2A1 as a Potential Diagnostic and Prognostic Biomarker for Sepsis | The study aimed to identify novel diagnostic and prognostic biomarkers related to monocytes and macrophages by using bulk RNA sequencing with integrated single-cell RNA sequencing. Differential expression analysis and weighted gene co-expression network analysis (WGCNA) were applied to detect sepsis and monocyte/macrophage-related genes. Least absolute shrinkage and selection operator (LASSO) and random forest regression analyses were used in combination to screen out prognostic genes. Single-cell RNA sequence profiling was utilized to further verify the expression of these genes on a single cell level. Receiver operating characteristic (ROC) curve and decision curve analysis (DCA) were also applied to verify the diagnostic value of the target biomarkers.<br>The study found that the intersections of the genes detected by differential expression and WGCNA analyses identified 141 overlapping candidate genes that were closely related to sepsis and macrophages. The LASSO and random forest regression analyses further screened out 17 prognostic genes. Single-cell RNA sequencing analysis detected that FCGR1A and BCL2A1 might be potential biomarkers for sepsis diagnosis and the diagnostic efficacy of BCL2A1 was further validated by ROC curve and DCA. | The study concludes that BCL2A1 had good diagnostic and prognostic value for sepsis, and that it can be applied as a potential and novel biomarker for the management of the disease. | 43 |
| A Transcriptomic Biomarker to Quantify Systemic Inflammation in Sepsis — A Prospective Multicenter Phase II Diagnostic Study | The study aimed to develop a dysregulated immune response biomarker that discriminates sepsis from uncomplicated infection. Marker candidates were screened by microarray and validated in a confirmation set of 246 medical and surgical patients. They identified up-regulated pathways reflecting innate effector mechanisms, while down-regulated pathways related to adaptive lymphocyte functions. A panel of markers composed of three up- (Toll-like receptor 5; Protectin; Clusterin) and 4 down-regulated transcripts (Fibrinogen-like 2; Interleukin-7 receptor; Major histocompatibility complex class II, DP alpha1; Carboxypeptidase, vitellogenic-like) described the magnitude of immune alterations.<br>The study found that the created gene expression score was significantly greater in patients with definite as well as with possible/probable infection than with no infection. Down-regulated lymphocyte markers were associated with prognosis with good sensitivity but limited specificity. | The study concludes that quantifying systemic inflammation by assessment of both pro- and anti-inflammatory innate and adaptive immune responses provides a novel option to identify patients-at-risk and may facilitate immune interventions in sepsis. | 45 |

**Table 3.** Diagnosis. Details the search results according to the chosen sepsis framework search term ‘Diagnosis’. * Literature Reference ** This paper appears in ‘Endotype’ and ‘Diagnosis’ searches

| Study | Methods and Results | Study Conclusion | Ref* |
| --- | --- | --- | --- |
| Integrated host-microbe plasma metagenomics for sepsis diagnosis in a prospective cohort of critically ill adults | Transcriptome profiling of colorectal tumors from patients with sepsis reveals an ethnic basis for viral infection risk and sepsis progression<br>The study conducted a retrospective transcriptomic analysis of colorectal cancer (CRC) tumors and adjacent non-tumor tissue from adult patients of Native Hawaiian and Japanese ethnicity who died from cancer-associated sepsis. The researchers examined differential gene expression in relation to patient survival and sepsis disease etiology.<br>The study found that Native Hawaiian CRC patients diagnosed with sepsis had a median survival of 5 months, compared to 117 months for Japanese patients. Transcriptomic analyses identified two distinct sepsis gene signatures classified as early response and late response sepsis genes that were significantly altered in the Native Hawaiian cohort. Analysis of canonical pathways revealed significant up and downregulation in mechanisms of viral exit from host cells and epithelial junction remodeling. | The study concludes that their transcriptomic approach advances understanding of sepsis heterogeneity by revealing a role of genetic background and defining patient subgroups with altered early and late biological responses to sepsis. Their findings may lead to personalized approaches in stratifying patient mortality risk for sepsis and in the development of effective targeted therapies for sepsis | 53 |
| Pre-symptomatic diagnosis of postoperative infection and sepsis using gene expression signatures | This study collected blood samples and clinical/laboratory data daily from 4385 patients undergoing elective surgery. An adjudication panel identified 154 patients with definite postoperative infection, of whom 98 developed sepsis. Transcriptomic profiling and subsequent RT-qPCR were undertaken on sequential blood samples taken postoperatively from these patients in the three days prior to the onset of symptoms. Comparison was made against postoperative day-, age-, sex- and procedure- matched patients who had an uncomplicated recovery (n =151) or postoperative inflammation without infection (n =148).<br>Specific gene signatures optimized to predict infection or sepsis in the three days prior to clinical presentation were identified in initial discovery cohorts. Subsequent classification using machine learning with cross-validation with separate patient cohorts and their matched controls gave high Area Under the Receiver Operator Curve (AUC) values. These allowed discrimination of infection from uncomplicated recovery (AUC 0.871), infectious from non-infectious systemic inflammation (0.897), sepsis from other postoperative presentations (0.843), and sepsis from uncomplicated infection (0.703). | Host biomarker signatures may be able to identify postoperative infection or sepsis up to three days in advance of clinical recognition. If validated in future studies, these signatures offer potential diagnostic utility for postoperative management of deteriorating or high-risk surgical patients and, potentially, other patient populations. | 54 |
| Validation of the Sepsis MetaScore for Diagnosis of Neonatal Sepsis | This study was designed as a secondary data analysis of cohorts from previously published studies. The authors searched for genome-wide expression studies of neonatal sepsis in PubMed, NCBI GEO, and EBI ArrayExpress. They included data sets only from studies of both neonates with sepsis and a reference/control class. For each study, the authors contacted the authors to gather laboratory data, including white blood cell (WBC) count, absolute neutrophil count (ANC), and C-reactive protein (CRP) level.<br>The authors found 3 cohorts with a total of 213 samples from control neonates and neonates with sepsis. The Sepsis MetaScore had an area under the receiver operating characteristic curve of 0.92–0.93 in all 3 cohorts. They also found that, as a diagnostic test for sepsis, it outperformed standard laboratory measurements alone and, when used in combination with another test(s), resulted in a significant net reclassification index (0.3–0.69) in 5 of 6 comparisons. | The Sepsis MetaScore had excellent diagnostic accuracy across 3 separate cohorts of neonates from 3 different countries. Further prospective targeted study will be needed before clinical application. | 57 |
| MicroRNAs combined with the TLR4/TDAG8 mRNAs and proinflammatory cytokines are biomarkers for the rapid diagnosis of sepsis. | This study included 40 patients with sepsis and 40 healthy controls. RNA-sequencing technology and bioinformatics analysis were applied to screen the DEMs between the two cohorts. The expression of these DEMs was subsequently verified by performing reverse transcription-quantitative PCR (RT-qPCR). In addition, IL-6, IL-21, C-X-C motif chemokine ligand-8 (CXCL8) and monocyte chemoattractant protein-1 (MCP-1) levels, along with T-cell death-associated gene 8 (TDAG8) and toll-like receptor 4 (TLR4) mRNA expression levels were assessed. The association between microRNA(miRNA/miR)-3663-3p and the secretion of various proinflammatory cytokines or TDAG8 and TLR4 mRNA expressions were subsequently evaluated by linear correlation analysis.<br>The results revealed 305 DEMs ($P < 0.05$; fold change $> 2$) between patients with sepsis and healthy controls. Among these, the top 18 up- and downregulated miRNAs were selected for RT-qPCR verification. In addition, the serum content of IL-6, IL-21, CXCL8 and MCP-1, and the expression of TDAG8 and TLR4 mRNAs were significantly increased in patients with sepsis compared with healthy controls. Moreover, in patients with sepsis, a positive correlation was identified between miR-3663-3p and the secretion of inflammatory cytokines or TDAG8 and TLR4 mRNA expression. A positive correlation was also elucidated between TDAG8 and TLR4 mRNA expression and proinflammatory cytokine/chemokine secretion. Receiver operating characteristic curve analysis of miR-3663-3p expression, IL-6, IL-21, CXCL8 and MCP-1 secretion and TDAG8 and TLR4 mRNA expression demonstrated that miRNA analysis may be invaluable for the diagnosis of sepsis. | The results determined that miR-3663-3p may be a potentially powerful diagnostic and predictive biomarker of sepsis and that the combined and simultaneous detection of several biomarkers, including proteins, miRNAs, and mRNA may be a reliable approach for the fast diagnosis and early identification of sepsis | 55 |
| Constructing a 10-core genes panel for diagnosis of pediatric sepsis | This study downloaded three sets of sepsis expression data (GSE13904, GSE25504, GSE26440) from GEO. Then, using the R limma package and WGCNA analysis to screen for core genes. Finally, the value of these core genes was confirmed by clinical samples.<br>Compared to normal samples, many abnormally expressed genes were obtained in the pediatric sepsis. WGCNA co-expression analysis showed that genes from blue and turquoise module were closely correlated with pediatric sepsis. The top 20 genes of the blue module of pediatric sepsis were mainly enriched in neutrophil degranulation, etc. The top 20 genes for turquoise module were mainly enriched in rRNA-containing ribonucleoprotein complexes exported from the nucleus, etc. The selected hub gene of pediatric sepsis was combined with the markers of cell surface and found 10 core genes (HCK, PRKCD, SIRPA, DOK3, ITGAM, LTB4R, MAPK14, MALT1, NLRC3, LCK). ROC showed that AUC of the 10 core genes for diagnosis of pediatric sepsis was above 0.9. | There were many abnormally expressed genes in patients with pediatric sepsis. The panel constructed by the 10 core genes was expected to become a biomarker panel for clinical application of pediatric sepsis. | 56 |
| Characterization of immune-related genes and immune infiltration features for early diagnosis, prognosis and recognition of immunosuppression in sepsis** | This study used a combination of machine learning approaches, including modified Lasso penalized regression and random forest (RF), to identify hub immune-related genes (IRGs) from multiple global datasets. The IRG score was generated through a linear combination of coefficients from logistic regression and the relative expression of each IRG. The diagnostic and prognostic values of the model were tested in independent validation cohorts from ArrayExpress databases. The authors also systematically correlated the IRG classifier with immunological characteristics from multiple perspectives, such as immune-related cells infiltrating, pivotal molecular pathways, and cytokine expression.<br>The study identified 8 hub IRGs (ADM, CX3CR1, DEFA4, HLA-DPA1, MAPK14, ORM1, RETN, and SLPI) that were combined to construct an IRG classifier. In the discovery cohort, the IRG classifier exhibited superior diagnostic efficacy and performed better in predicting mortality than clinical characteristics or MARS/SRS endotypes. Encouragingly, similar results were observed in the ArrayExpress databases. The use of hydrocortisone in the IRG high-risk subgroup was associated with increased risk of mortality. In the IRG low-risk phenotypes, NK cells, T helper cells, and infiltrating lymphocyte (IL) were significantly richer, while T cells regulatory (Tregs) and myeloid-derived suppressor cells (MDSC) were more abundant in the IRG high-risk phenotypes. | The study established a diagnostic and prognostic model based on 8 IRGs that is closely correlated with responses to hydrocortisone and immunosuppression status. This model might facilitate personalized counseling for specific therapy. The results suggest that components of the immune system play a crucial role in the occurrence and development of sepsis. | 37 |

**Table 4.** Sepsis progression. Details the search results according to the chosen sepsis framework search term ‘Progression’. * Literature Reference

| Study | Methods and Results | Conclusion | Ref* |
| --- | --- | --- | --- |
| Host Gene Expression to Predict Sepsis Progression. | This study aimed to develop transcriptomic models to predict progression to sepsis or shock within 72 hours of hospitalization and to validate previously identified transcriptomic signatures in the prediction of 28-day mortality. The authors hypothesized that host gene expression could predict sepsis progression and mortality. The authors used RNA sequencing of whole blood from 277 patients with infection, sepsis, or septic shock enrolled at four hospitals. They performed differential gene expression analysis and predictive modeling to identify signatures for sepsis progression and mortality.<br>The authors found no differential gene expression or predictive signature for sepsis progression defined by the Sepsis-3 category. However, they found differential gene expression and moderate predictive ability for sepsis progression defined by new organ dysfunction or ICU admission/mortality. They also validated four previously published gene signatures for sepsis mortality. | The authors concluded that host gene expression was unable to predict sepsis progression by the Sepsis-3 category, but could predict more severe outcomes<sup>3</sup>[3]. They suggested that transcriptomic methods should account for biological and clinical heterogeneity in sepsis. | 58 |
| "Transcriptome profiling of colorectal tumors from patients with sepsis reveals an ethnic basis for viral infection risk and sepsis progression" | The study conducted a retrospective transcriptomic analysis of colorectal cancer (CRC) tumors and adjacent non-tumor tissue from adult patients of Native Hawaiian and Japanese ethnicity who died from cancer-associated sepsis. The researchers examined differential gene expression in relation to patient survival and sepsis disease etiology.<br>The study found that Native Hawaiian CRC patients diagnosed with sepsis had a median survival of 5 months, compared to 117 months for Japanese patients. Transcriptomic analyses identified two distinct sepsis gene signatures classified as early response and late response sepsis genes significantly altered in the Native Hawaiian cohort. Analysis of canonical pathways revealed significant up and downregulation in mechanisms of viral exit from host cells and epithelial junction remodeling. | The study concludes that their transcriptomic approach advances the understanding of sepsis heterogeneity by revealing the role of genetic background and defining patient subgroups with altered early and late biological responses to sepsis. Their findings may lead to personalized approaches in stratifying patient mortality risk for sepsis and in the development of effective targeted therapies for sepsis. | 59 |

**Table 5.** Sepsis Severity. Details the search results according to the chosen sepsis framework search term ‘Severity’. * Literature Reference

| Study | Method and Results | Study Conclusion | Ref* |
| --- | --- | --- | --- |
| "Predicting sepsis severity at first clinical presentation: The role of endotypes and mechanistic signatures" | The study aimed to identify gene expression signatures that predict subsequent severity in sepsis patients at first clinical presentation. Blood RNA-Seq and clinical data were collected from 348 patients in four emergency rooms (ER) and one intensive-care-unit (ICU), and 44 healthy controls. Gene expression profiles were analyzed using machine learning and data mining to identify clinically relevant gene signatures reflecting disease severity, organ dysfunction, mortality, and specific endotypes/mechanisms.<br>**\*\*Results:\*\*** The study found that gene expression signatures were obtained that predicted severity/organ dysfunction and mortality in both ER and ICU patients with accuracy/AUC of 77-80%. Network analysis revealed these signatures formed a coherent biological program, with specific but overlapping mechanisms/pathways. Patients with early sepsis could be stratified into five distinct and novel mechanistic endotypes, named Neutrophilic-Suppressive/NPS, Inflammatory/INF, Innate-Host-Defense/IHD, Interferon/IFN, and Adaptive/ADA, each based on approximately 200 unique gene expression differences, and distinct pathways/mechanisms. Endotypes had varying overall severity with two severe (NPS/INF) and one relatively benign (ADA) groupings. A 40 gene-classification tool (accuracy=96%) and several gene-pairs (accuracy=89-97%) accurately predicted endotype status in both ER and ICU validation cohorts. | **** [Duplicate with endotype category]*****<br>**\*\*Conclusions:\*\*** The study concludes that the severity and endotype signatures indicate that distinct immune signatures precede the onset of severe sepsis and lethality, providing a method to triage early sepsis patients. | 29 |
| Estimates of gene ensemble noise highlight critical pathways and predict disease severity in H1N1, COVID-19 and mortality in sepsis patients | This study developed an alternative approach to dissect disease-associated molecular changes by defining gene ensemble noise as a measure that represents a variance for a collection of genes encoding for either members of known biological pathways or subunits of annotated protein complexes and calculated within an individual. The gene ensemble noise allows for the holistic identification and interpretation of gene expression disbalance on the level of gene networks and systems. By comparing gene expression data from COVID-19, H1N1, and sepsis patients, the study identified common disturbances in a number of pathways and protein complexes relevant to the sepsis pathology.<br>The study found that among others, these include the mitochondrial respiratory chain complex I and peroxisomes. This suggests a Warburg effect and oxidative stress as common hallmarks of the immune host-pathogen response. Finally, it was shown that gene ensemble noise could successfully be applied for the prediction of clinical outcome namely, the mortality of patients. | The study concludes that gene ensemble noise represents a promising approach for the investigation of molecular mechanisms of pathology through a prism of alterations in the coherent expression of gene circuits. | 60 |

**Table 6.** Benchmark. Details the search results according to the chosen sepsis framework search term ‘Benchmark’. * Literature Reference

| Study | Method and Results | Conclusion | Ref* |
| --- | --- | --- | --- |
| Benchmarking sepsis gene expression diagnostics using public data. | A systematic search for publicly available gene expression data in sepsis. The authors tested each gene expression classifier in all included datasets and created a public repository of sepsis gene expression data to encourage their future re-use.<br>Several new gene expression classifiers have been recently published for better sepsis diagnostics, including an 11-gene 'Sepsis MetaScore', the FAIM3:PLAC8 ratio, and the Septicyte Lab. The authors performed a systematic search for publicly available gene expression data in sepsis and tested each gene expression classifier in all included datasets. They also created a public repository of sepsis gene expression data to encourage their future re-use. | Three diagnostics do not show significant differences in overall ability to distinguish non-infectious SIRS from sepsis, though the performance in some datasets was low (AUC<0.7) for the FAIM3:PLAC8 ratio and Septicyte lab. The Septicyte Lab also demonstrated significantly worse performance in discriminating infections as compared to healthy controls. Overall, public gene expression data is a useful tool for benchmarking gene expression diagnostics. | <sup>63</sup> |
| The leukocyte non-coding RNA landscape in critically ill patients with sepsis | The authors obtained whole blood from 156 patients with sepsis and 82 healthy subjects among whom eight were challenged with lipopolysaccharide in a clinically controlled setting (human endotoxemia). They performed next-generation microarray analysis of leukocyte RNA to evaluate protein-coding, long and small non-coding RNA expression.<br>The authors found that long non-coding RNA and, to a lesser extent, small non-coding RNA were significantly altered in sepsis relative to health. Long non-coding RNA expression, but not small non-coding RNA, was largely recapitulated in human endotoxemia. Integrating RNA profiles and plasma protein levels revealed known as well as previously unobserved pathways, including non-sensory olfactory receptor activity. | The transcriptional changes in critically ill patients with sepsis are not exclusive to protein-coding RNAs. Whole blood long non-coding RNAs, and to a lesser extent small non-coding RNAs, were significantly altered in sepsis patients relative to healthy subjects. The pattern of protein-coding and long non-coding RNA profiles in sepsis was mimicked by expression profiles in a human endotoxemia model. This study provides a benchmark dissection of the blood leukocyte 'regulome' that can facilitate prioritization of future functional studies. | <sup>64</sup> |

**Supplement Table 7:**
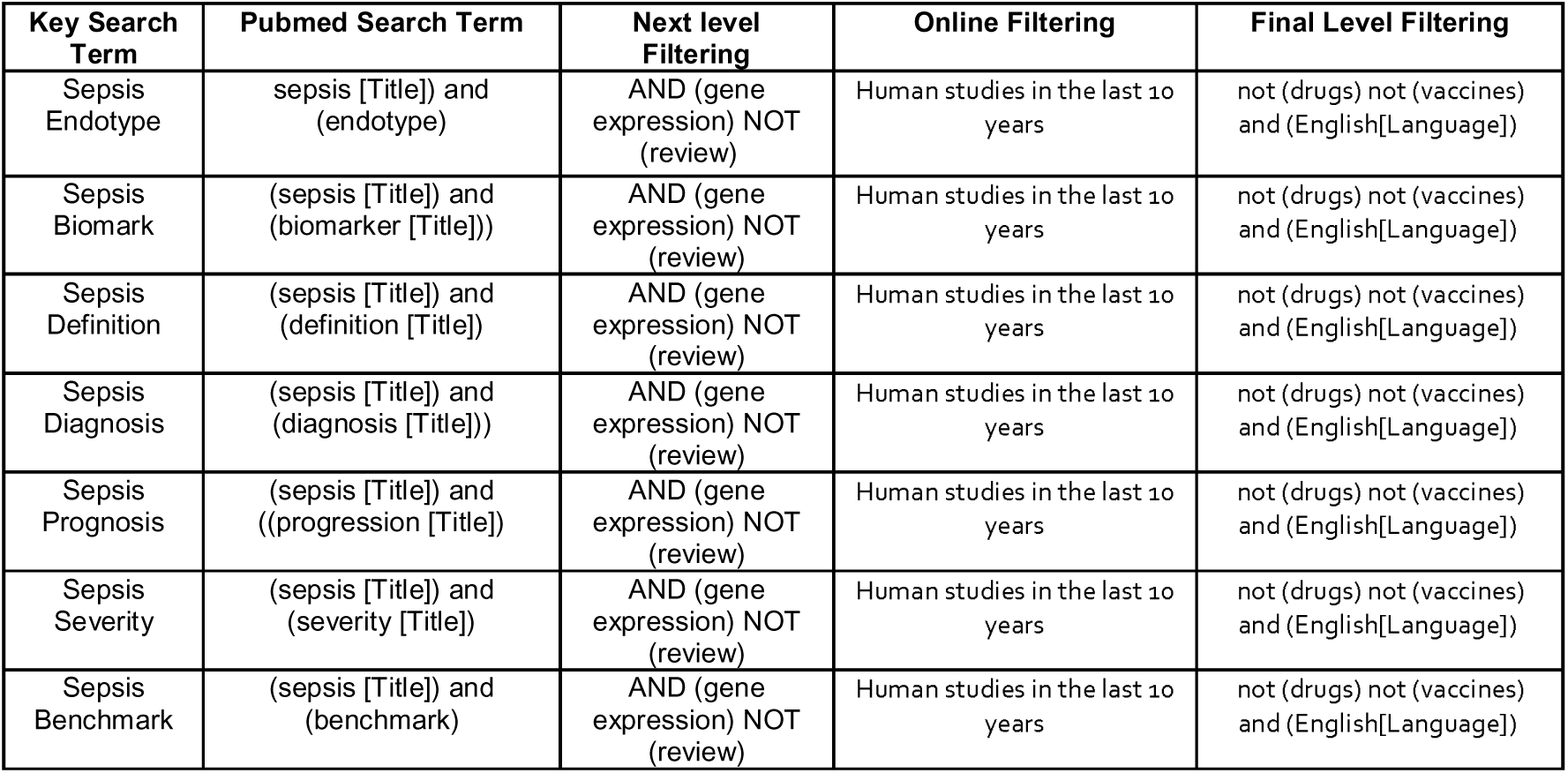
Pubmed Search Strategies. Table shows the different level of filtering applied to the different search terms. For sepsis Endotype and Benchmark the search terms were expanded to include the full body of the text. This was in order to expand the search and ensure inclusion of all relevant inform

